# Direct observation of repeated infections with endemic coronaviruses

**DOI:** 10.1101/2020.04.27.20082032

**Authors:** Marta Galanti, Jeffrey Shaman

**Author notes:** Corresponding author - contact information, Dr. Marta Galanti, Post-Doctoral Research Scientist, Department of Environmental Health Sciences, Mailman School of Public Health, 722 West 168^th^ St. NY, NY 10032, Columbia University.

## Abstract

**Background:** While the mechanisms of adaptive immunity to pandemic coronavirus SARS-CoV-2 are still unknown, the immune response to the widespread endemic coronaviruses HKU1, 229E, NL63 and OC43 provide a useful reference for understanding repeat infection risk.

**Methods:** Here we used data from proactive sampling carried out in New York City from fall 2016 to spring 2018. We combined weekly nasal swab collection with self-reports of respiratory symptoms from 191 participants to investigate the profile of recurring infections with endemic coronaviruses.

**Results:** During the study, 12 individuals tested positive multiple times for the same coronavirus. We found no significant difference between the probability of testing positive at least once and the probability of a recurrence for the beta-coronaviruses HKU1 and OC43 at 34 weeks after enrollment/first infection. We also found no significant association between repeat infections and symptom severity but strong association between symptom severity and belonging to the same family.

**Conclusion:** This study provides evidence that re-infections with the same endemic coronavirus are not atypical in a time window shorter than 1 year and that the genetic basis of innate immune response may be a greater determinant of infection severity than immune memory acquired after a previous infection.

## Background

The new coronavirus SARS-CoV-2 appears to have emerged in humans in the Hubei province of China during November 2019 [1]. Human to human transmission was confirmed in early January, and since then the virus has rapidly spread to all continents. The outbreak was declared a pandemic by the WHO on March 11th. As of April 10th, it had spread to over 180 countries with 1,521,252 confirmed cases and 92,798 deaths reported [2].

Symptoms associated with SARS-CoV-2 vary from none to extremely severe, with elder adults and people with underlying medical conditions more at risk for developing severe and potentially fatal disease [3]. At present, there is no vaccine or approved antiviral treatment for SARS-CoV-2, and therapies rely principally on symptom management. Many institutions across the world are working to develop a SARS-CoV-2 vaccine, and clinical trials with some vaccine candidates have already begun [4].

As the pandemic progresses, infecting millions of people across the world, a key question is whether individuals upon recovery are prone to repeat infection. A recent animal challenge study showed evidence of (at least) short-term protection against re-infections in rhesus macaques experimentally re-infected 4 weeks after first infection [5]. Typically, infections by different viruses trigger different adaptive immune responses: viruses like measles elicit life-long immunity; whereas others, like influenza, do not. Two main processes appear to be responsible for the short-lived immunity engendered against some pathogens: 1) waning of antibodies and memory cells in the host system; and 2) antigenic drift of the pathogen that enables escape from the immunity built against previous strains.

To contextualize the issue of protective immunity to SARS-CoV-2, we here present findings from a recent proactive sampling project carried out in New York City (NYC) that documented rates of infection and re-infection among individuals shedding seasonal CoV (types: HKU1, 229E, NL63 and OC43). The results are discussed and analyzed in the broader context of coronavirus infections.

## Methods

Data are derived from sampling performed between October 2016 and April 2018 as part of the Virome project, a proactive sampling of respiratory virus infection rates, associated symptom self-reports and rates of seeking clinical care. We enrolled 214 healthy individuals from multiple locations in the Manhattan borough of New York City. Cohort composition is described in [6] and includes: children attending two daycares, along with their siblings and parents; teenagers and teachers from a high school; adults working at two emergency departments (a pediatric and an adult hospital); and adults working at a university medical center. The cohort was obtained using convenience sampling, and all participants were younger than 65 years. While the study period spanned 19 months from October 2016 to April 2018, some individuals enrolled for a single cold and flu season (October - April) and others for the entire study period. Participants (or their guardians, if minors) provided informed consent after reading a detailed description of the study (CUMC IRB AAAQ4358).

Nasopharyngeal samples were collected by study coordinators once a week irrespective of participant symptoms. Samples were screened using the GenMark eSensor RVP system for 18 different respiratory viruses, including coronavirus 229E, NL63, OC43, and HKU1. Sample collection and extraction followed the same protocol as in [7].

In addition, participants completed daily self-reports rating nine respiratory illness-related symptoms (fever, chills, muscle pain, watery eyes, runny nose, sneezing, sore throat, cough, chest pain), each of which was recorded on a Likert scale (0=none, 1=mild, 2=moderate, 3=severe), see [6] for further survey details.

For this analysis, only the 191 participants who contributed at least six separate pairs of nasopharyngeal samples in the same season were included. We defined an infection (or viral) episode as a group of consecutive weekly specimens from a given individual that were positive for the same virus (allowing for a one-week gap to account for false negatives and temporary low shedding). We classified all infection episodes as symptomatic or asymptomatic according to individual symptom scores in the days surrounding the date of the first positive swab of an episode. We used multiple definitions as a standard for symptomatic infection does not exist (Table 1). These symptom definitions are described in reference to a −3 to +7-day window around the date of the initial positive swab for each infection episode. The daily symptom score is defined as the sum of the 9 individual symptoms (range: 0–27) on a given day. Total symptom score is the daily symptom score summed over the −3 to +7-day window.

**Table 1.**
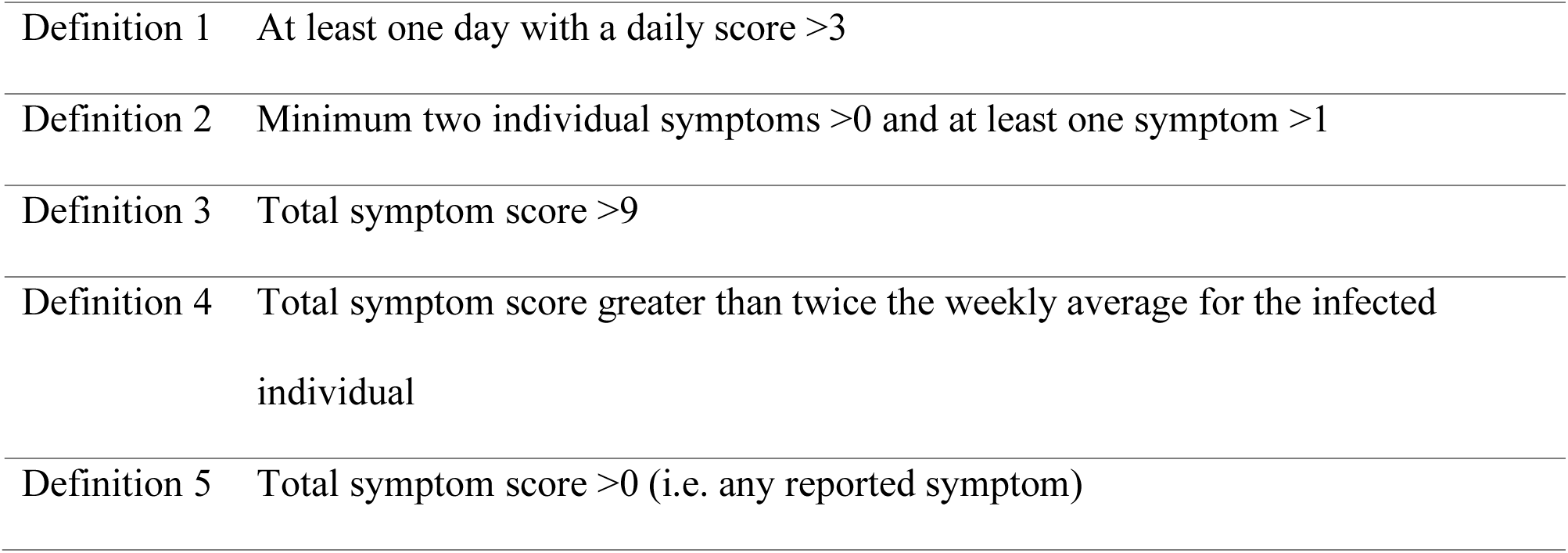
Definitions of symptomatic infections. All symptom definitions are described in reference to a −3/+7 days window around the date of the initial positive swab for an infection episode. Note, Definition 4 is relative to an individual’s long-term average total symptom score.

|  |  |
| --- | --- |
| Definition 1 | At least one day with a daily score >3 |
| Definition 2 | Minimum two individual symptoms >0 and at least one symptom >1 |
| Definition 3 | Total symptom score >9 |
| Definition 4 | Total symptom score greater than twice the weekly average for the infected individual |
| Definition 5 | Total symptom score >0 (i.e. any reported symptom) |

We used Survival Analysis methods to estimate the probability of infection (as a function of time from enrollment) and the waning of protective immunity following first infection for each type of coronavirus. Specifically, we used the Kaplan Meier estimator S(t) to estimate 1) the probability of being infected with each coronavirus type and 2) the probability of being reinfected with the same coronavirus type following a previous documented infection. *I*(*t*) measures the probability of having tested positive for a given coronavirus type by time t:

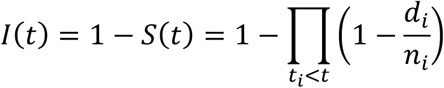

Time *t* is measured in weeks from enrollment in the first analysis and from the previous documented infection with a specific coronavirus type in the second analysis; *d_i_*, are the participants testing positive *i* weeks after enrollment (after first infection) and *n_i_* are the participants that are still enrolled *i* weeks after enrollment (after first infection). The denominator *n_i_*, corrects for participants withdrawing from the study at different time by right censoring.

The estimators for the probability of infection and reinfection are compared statistically using the log rank test. We used Fisher’s exact test to analyze the difference between symptoms developed during subsequent infections and ANOVA comparison to test differences in symptom scores reported by different family clusters. We restricted the last analysis to the family clusters within the cohort that presented at least 3 coronavirus infections during the study.

## Results

Among all participants enrolled, 86 individuals tested positives at least once during the study for any coronavirus infection. 48 individuals tested positive at least once for OC43, 31 tested positive for 229E, 15 tested positive for NL63 and 28 tested positive for HKU1. Figure 1 shows a Kaplan-Meier plot estimating the probability of becoming infected with each coronavirus within x weeks following enrollment (see Supplementary Table S1 for the number of individuals infected and censored at each time point). OC43 was the most widely diffused virus: the probability of testing positive following 80 weeks in the study was 0.47. In contrast, NL63 was the least frequently isolated coronavirus type: the probability of testing positive after 80 weeks was 0.17. Among the study participants, 12 individuals tested positive multiple times during the study for the same coronavirus: 9 tested positive multiple times for OC43, 2 tested positive twice for HKU1, 1 tested positive twice for 229E and nobody tested positive multiple times for NL63. Among the 9 participants with multiple OC43 infections, 3 individuals experienced 3 separate infection episodes, and the other 6 experienced 2 separate episodes. The median time between reinfection events was 37 weeks. The shortest time for a reoccurrence of infection was 4 weeks (OC43), the longest was 48 weeks (OC43). Among the 12 individuals testing positive multiple times for the same coronavirus, 9 were children aged between 1 and 9 years at enrollment, and 3 were adults aged between 25 and 34 years (see Supplementary Table S2 for characteristics of the repeated infections).

**Figure 1:**
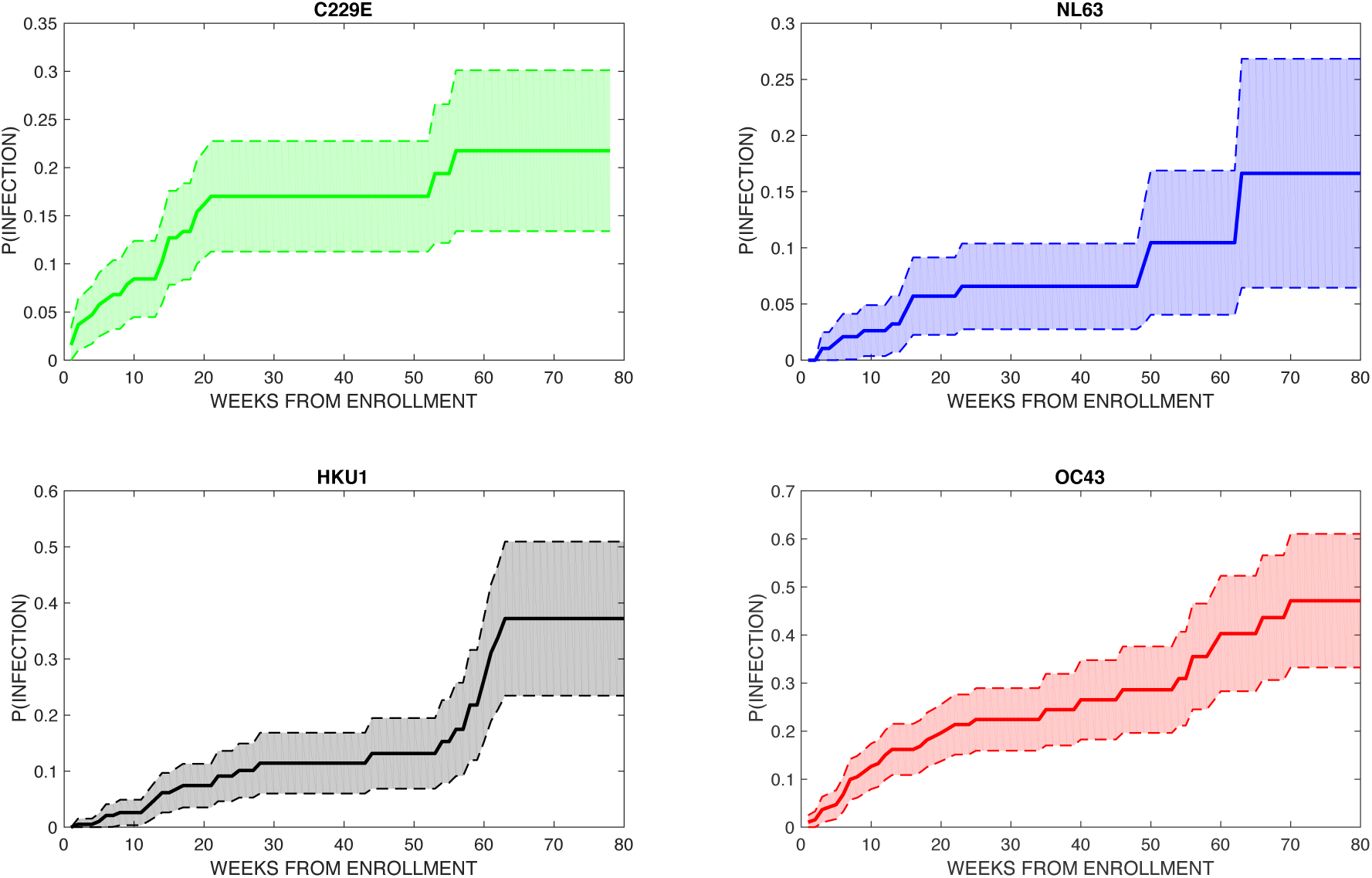
Kaplan-Meier plots showing the probability of testing positive within *x* weeks after enrollment for each of the 4 types of seasonal coronavirus. The shaded area is the 95% CI.

Figure 2 shows a Kaplan-Meier plot estimating the probability of becoming re-infected with the same beta-coronavirus (OC43 and HKU1) within x weeks after a previously documented infection (see Supplementary Table S3 for the number of individuals infected and censored at each time point). A comparison between the data shown in Fig 2 and Fig 1 finds no significant differences between the probability of testing positive at least once and the probability of a recurrence for both HKU1and OC43 at 34 weeks after enrollment/first infection.

**Figure 2:**
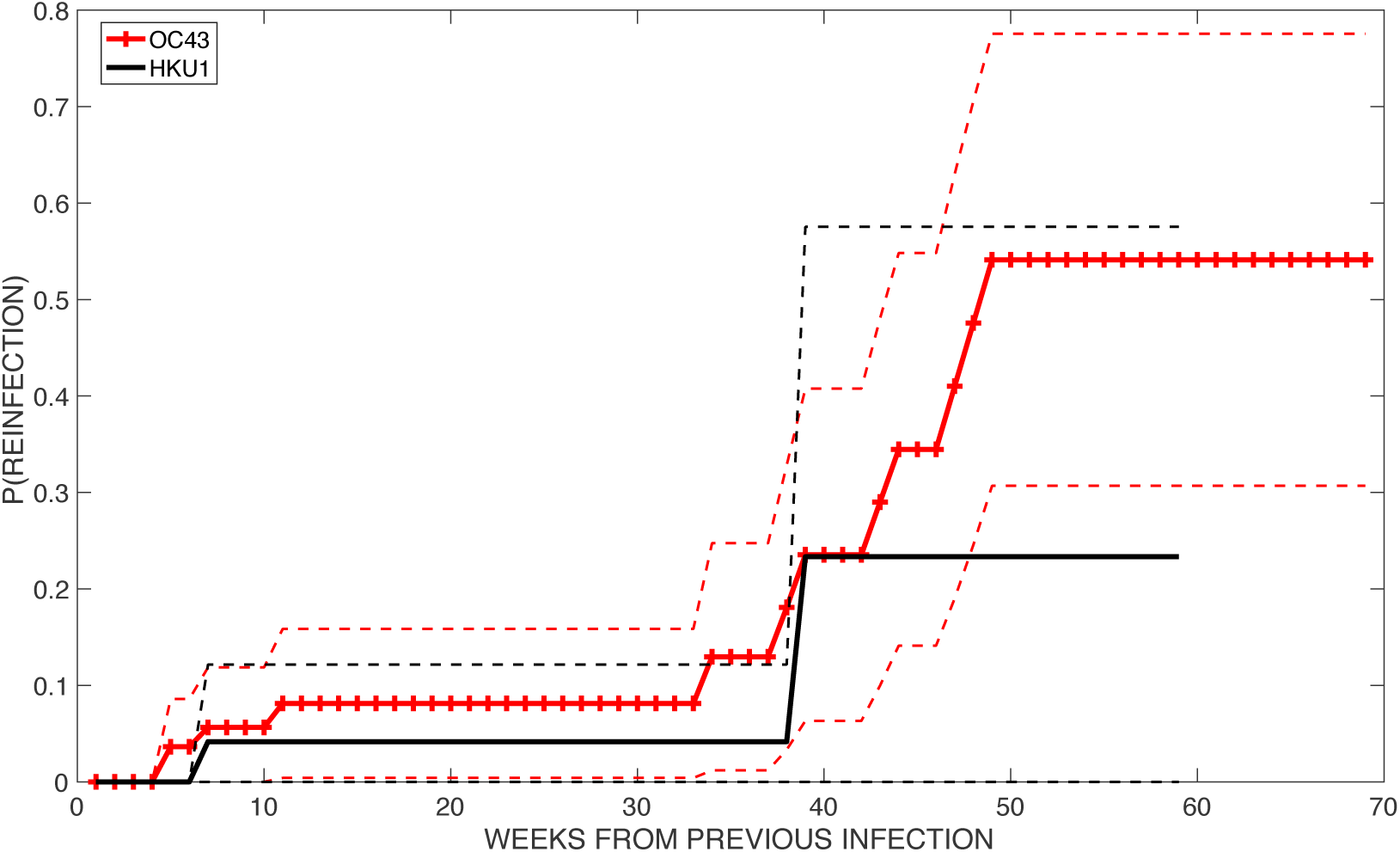
Probability of becoming re-infected with the same beta-coronavirus type (OC43 in red crossed line and HKU1 in black straight line) within x weeks after a first documented infection. Dashed lines show the 95% CI.

To control for false positive PCR results, we tested the sensitivity of the findings to different choices of the positivity threshold used in RVP testing (see Supplementary Text 1 and Supplementary Figures S1 toS 4). The probability of reinfection with beta-coronaviruses at > 38 weeks after prior infection was robust across different thresholds, whereas short terms reinfection signals could be an artifact due to PCR amplification. This shifted threshold also yields a statistically significant difference between the probability of testing positive at least once and the probability of a recurrence after first infection until week 43 (*p* = 0.04).

There was no significant difference in the likelihood of experiencing symptomatic infection between the first and subsequent infection episodes by any of the 5 definitions provided in Table 1. In particular, all the individuals who were completely asymptomatic during the first recorded occurrence, did not report any symptoms during subsequent infection(s) with the same coronavirus type. However, there was a significant association between severity of symptoms associated with any coronavirus infection and belonging to the same family cluster (*p*<.0001, one-way analysis of variance). Figure 3 shows the total symptom score associated with any coronavirus infection for infections grouped by family cluster.

**Figure 3:**
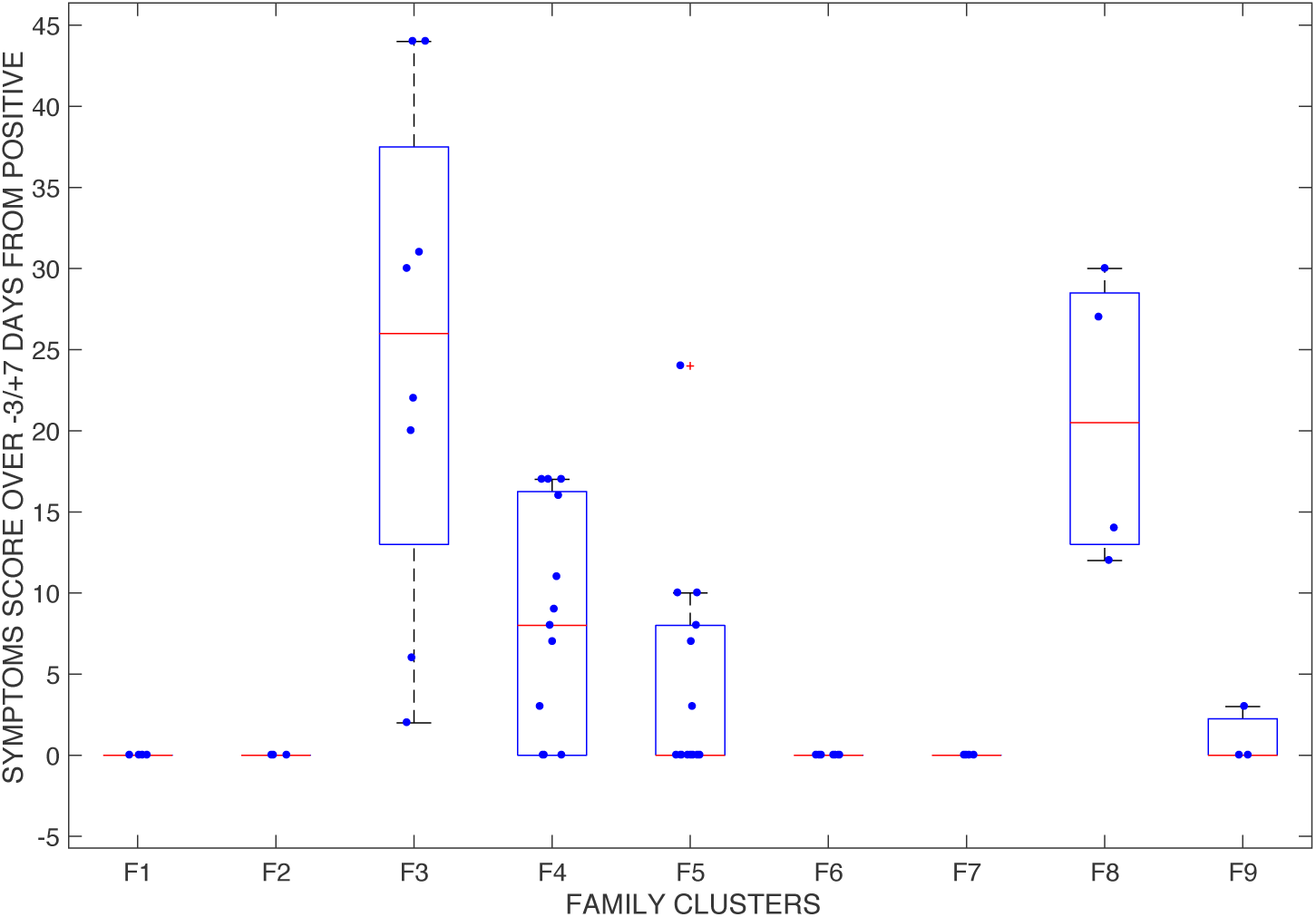
Total symptom score associated with infections by any coronavirus type. Each point represents an infection event, and each cluster represents a family group. Each family group F1 to F9 is composed of a parent and 1 to 4 children.

## Discussion

As the SARS-CoV-2 pandemic spreads to millions of individuals worldwide, it is extremely important to understand the mechanisms of protective immunity elicited by infection. Until direct observations of adaptive immune response to SARS-CoV-2 become available, analyses of protective immunity elicited by other coronaviruses may offer useful insights.

Several studies in the last four decades have shown that infections with the 4 endemic coronaviruses 229E, OC43, NL63 and HKU are common in the general population [8] [9]. Infection with these viruses generally produces mild and even asymptomatic infection [10]. Serological studies have shown that more than 90% of the population presents a baseline level of antibodies against these endemic coronaviruses, with first seroconversion occurring at a young age [11] [8]. Shortly after infection, baseline antibody titers increase sharply; this response has been demonstrated for both natural and experimentally-induced infections [12] [13] [9].

Antibody titers start increasing roughly one week following infection, reach a peak after about 2 weeks [13], and by 4 months to 1 year have returned to baseline levels [13] [9]. A challenge study [13] showed that the likelihood of developing an infection after inoculation correlated with participants’ concentration of antibodies at enrollment. Moreover, a positive correlation has been shown between antibody rise after infection, severity of clinical manifestation and viral shedding [12], with milder cases linked to less substantial post-infection antibody rises.

Instances of natural re-infections with the same virus type have been documented previously [9] in which repeated infections with OC43 and 229E were recorded by serological testing. Subsequent infections were separated by at least 8 months, though study participants were tested every 4 months. Participants in a separate challenge study were inoculated with coronavirus 229E and then re-challenged with the same virus after one year [13]. In most cases, re-infection occurred, though it presented with decreased symptoms severity and shortened duration of shedding.

The adaptive immune response to coronavirus is mainly directed towards the most variable part of the virus, a region that is not conserved across types; consequently, cross-reactive protection between different types does not appear to be an important factor [14, 15]. In addition, the effects of antigenic drift on re-infection have not been elucidated [16] and more studies are warranted to understand whether repeat infections are ascribable to rapid virus evolution rather than a decline in antibody titers.

The mild pathogenicity of seasonal coronavirus infection (with immune response often restricted to the upper respiratory trait) is also often regarded as the reason for short-lived immunity. Coronavirus infections, and the adaptive immunity acquired towards them, have also been studied in animals. In a study on porcine respiratory coronavirus (PRCV), which causes subclinical infections in pigs, antibody titers waned approximately one year after experimental infection [17]. In contrast, an experimental study on murine coronavirus (MHV), which produces severe, systemic infections in mice, has shown an interplay between virus-specific antibodies and T cells, that upon survival in the host lead to life-long protection against reinfection [18]. Similarly, a longer immunity profile has been hypothesized for SARS and MERS due to their increased severity and to the systemic response that infection induces [14]. Specific antibodies were detectable for at least 2 years in SARS and MERS survivors [19] [20]. Although longitudinal studies on SARS survivors have not detected specific SARS IGG antibody persistence 5 years after infection, they have found that specific memory T cells persist in the peripheral blood of recovered SARS patients, and at higher levels in patients who experienced severe disease [21]. Whether the presence of these memory T cells would be enough to induce a fast, protective response upon reinfection with SARS has not been assessed.

Our study confirms that seasonal coronaviruses are widespread in the general population with infections directly documented for a large fraction of the participants in our study. The methods for our analysis are based on the hypothesis that infection probabilities are comparable among participants enrolled at different times in the study. However, the seasonality of endemic coronaviruses, which are mostly absent during the summer months, and the relative magnitude across years of seasonal coronavirus epidemics are limitations. In US the prevalence of OC43 during the 2016–17 season was much higher than during the 2017–18 season, whereas the opposite trend was observed for HKU1 [22]. Moreover, our estimates of infection and re-infection probabilities must be considered as a lower bound, due to the occurrence of weekly swabs missed by the participants and due to the design of the study itself, which may have missed infections of short duration in between consecutive weekly tests. Nevertheless, this study confirms that re-infections with the same coronavirus type occur in a time window shorter than 1 year, and finds no significant association between repeat infections and symptom severity. Instead, it provides evidence of possible genetic determinants of innate immune response, as individuals asymptomatic during first infection did not experience symptoms during subsequent infections, and members of the same families reported similar symptom severity. We recognize that the self-reporting of symptoms is an important limitation in this analysis and that parents reported symptoms for their dependents, which possibly introduced bias. Moreover, the majority of the repeated coronavirus infections were found in children, a cohort more vulnerable to infection because of their immature immune system [23], and 26% of the episodes in the repeated infections were co-infections with other respiratory viruses (see Supplementary Table S2). Another potential limitation of our study is the high sensitivity of PCR tests, that can amplify very small amounts of genetic material, possibly not ascribable to active infections. However, the occurrence of repeated infections separated by at least 38 weeks, was corroborated by repeating the analysis with different positivity thresholds for the RVP.

More studies analyzing the genetic basis of individual response to coronavirus infections are warranted. Even though the endemic coronaviruses are very rarely associated with severe disease, their widespread diffusion together with the fact that OC43 and HKU1 belong to the same beta-coronavirus genus as SARS-CoV2 offer important opportunities for investigation.

## Data Availability

Data are reported in the supplementary material.

